# A Novel Therapeutic Mechanism for Nicotine Craving in Schizophrenia

**DOI:** 10.64898/2026.03.14.26348404

**Authors:** Heather Burrell Ward, Jillian G. Connolly, Sophia H. Blyth, Simon Vandekar, Baxter P. Rogers, Mark A. Halko, Catie Chang, Hilary Tindle, L. Elliot Hong, A. Eden Evins, Stephan Heckers, Roscoe O. Brady

**Author notes:** <u>Corresponding Author</u> Heather Burrell Ward, MD 1601 23^rd^ Ave S Nashville, TN 37212.

## Abstract

**Objective:** Tobacco use is a leading cause of mortality in schizophrenia, but treatments are partially effective. Default mode network (DMN) pathology is linked to tobacco use in schizophrenia, and transcranial magnetic stimulation (TMS) applied to the DMN affects craving in schizophrenia. To advance TMS therapeutics for tobacco use in schizophrenia, we used TMS experiments to 1) determine optimal stimulation parameters then 2) compare our optimal parameters against a well-established, effective TMS intervention for craving.

**Methods:** In Protocol Optimization TMS, nicotine-using individuals with schizophrenia (n=10) received single sessions of DMN-targeted TMS with pre/post neuroimaging and craving assessment. Neuroimaging analysis revealed bilateral parietal DMN connectivity was associated with craving change. In Comparative Effectiveness TMS (n=62), nicotine-using individuals with schizophrenia and non-psychosis controls participated in a crossover study comparing DMN-targeted and left dorsolateral prefrontal cortex (DLFPC)-targeted TMS with pre/post neuroimaging and craving assessment. Mixed effects models were used to determine effects of target, group, and relationship between craving change and connectivity change.

**Results:** In Protocol Optimization TMS, increased craving was associated with increased bilateral parietal DMN connectivity (mean pFDR<0.012, r=0.60). In Comparative Effectiveness TMS, both interventions reduced craving (DLPFC: p=0.0015; DMN: p=0.0054) and bilateral parietal DMN connectivity (DLPFC: p=0.024; DMN: p=0.022). There was an interaction of bilateral parietal DMN connectivity change, group, and age (p=0.001) where connectivity change was associated with craving change in older individuals with schizophrenia (p=0.041) but not other groups.

**Conclusions:** Bilateral parietal DMN connectivity is a novel mechanism underlying craving in schizophrenia that can be engaged for therapeutic benefit.

## Introduction

Tobacco use is the top preventable cause of mortality in schizophrenia. The prevalence of tobacco use in schizophrenia is over 60% (1, 2), which contributes to a 20-year decreased life expectancy compared to the general population (3). The underlying pathophysiology of nicotine dependence in schizophrenia remains unknown, and current treatments are inadequate.

Smoking cessation interventions for schizophrenia are derived from non-psychosis populations. Both pharmacologic and neuromodulation treatments for nicotine use are less effective for people with schizophrenia (4). This suggests interventions targeting networks derived from people without psychosis may not treat pathophysiology underlying nicotine dependence in schizophrenia. Transcranial magnetic stimulation (TMS) is cleared by the U.S. Food and Drug Administration (FDA) for smoking cessation in the general population but has mixed results in schizophrenia (5).

These mixed results suggest there may be additional brain circuitry contributing to nicotine use in schizophrenia. Previous work has identified schizophrenia-specific links between nicotine use and default mode network (DMN) organization. We observed that individual variation in the severity of nicotine use was linked to individual variation in parietal DMN organization such that heavier smokers had pathological DMN organization (6). This relationship was *specific* to schizophrenia and not observed in smokers without schizophrenia.

DMN connectivity has been previously linked to nicotine use. Higher DMN connectivity is linked to higher nicotine craving in controls and schizophrenia (7, 8). DMN hyperconnectivity is also a well-established finding in schizophrenia (9). Acute nicotine administration *normalizes* DMN hyperconnectivity in people with schizophrenia, but not in controls (6). These results suggest that nicotine normalizes DMN dysfunction in schizophrenia, so modulating DMN connectivity may be a novel target for nicotine craving in schizophrenia.

To test if modulating the DMN would affect nicotine craving in schizophrenia, we compared stimulation parameters in Protocol Optimization TMS using single TMS sessions applied to a personalized left parietal DMN target and showed that DMN-targeted TMS affected craving (Figure 1A) (10). To understand the brain mechanism underlying this craving change, in the current analysis, we used an unrestricted brain-wide analysis to understand how connectivity from the left parietal TMS target was related to craving change. We observed that reduced craving was related to reduced connectivity from the left parietal TMS target to a right parietal DMN region and that continuous theta burst (cTBS) most effectively reduced connectivity. This finding showed that craving change was the result of bilateral DMN connectivity change and that cTBS moved connectivity in the therapeutic direction.

**Figure 1.**
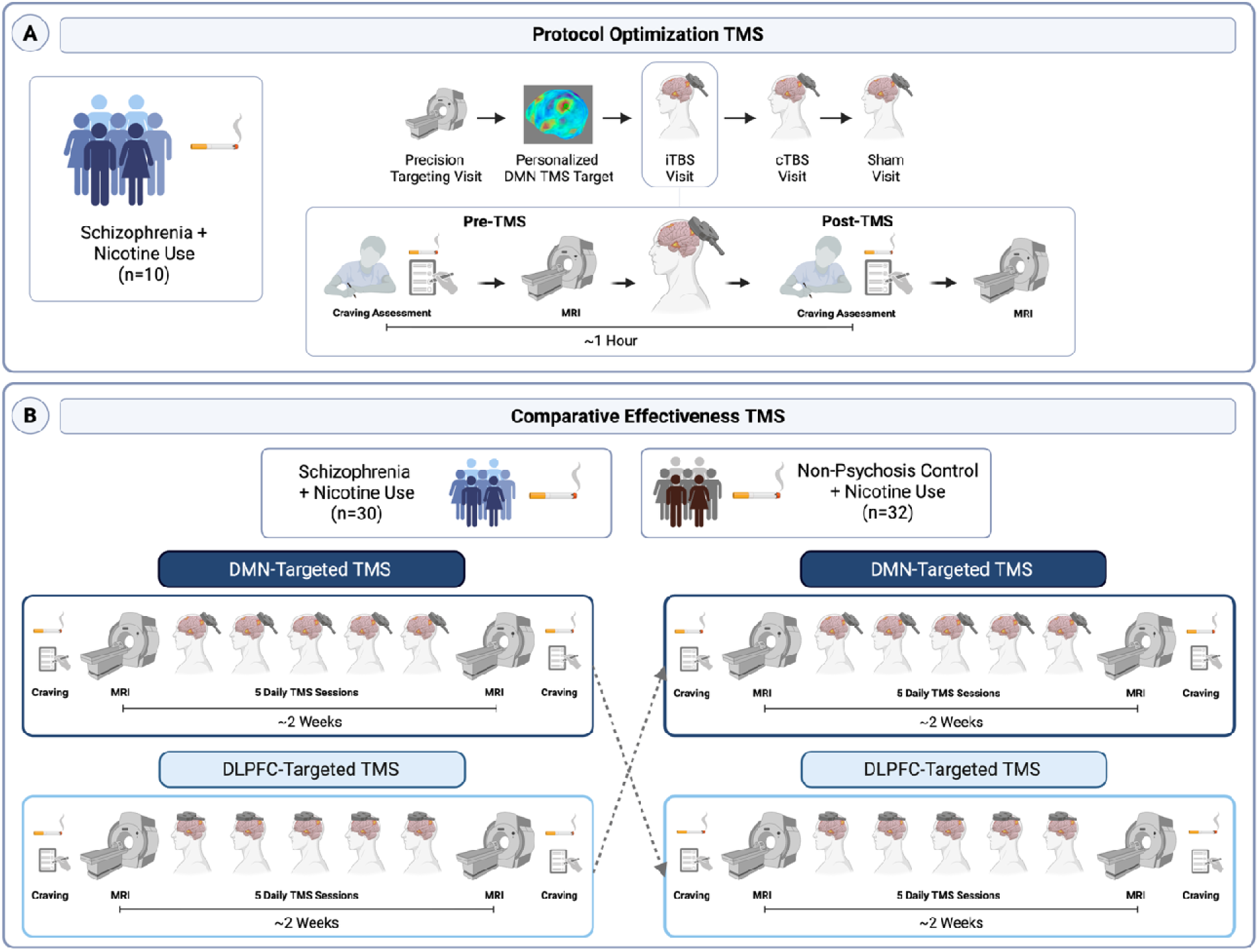
Study Designs. We tested single- and multiple-sessions of default mode network (DMN)-targeted transcranial magnetic stimulation (TMS) to understand the brain mechanism underlying craving in schizophrenia. In the Protocol Optimization TMS study (Figure 1A), 10 individuals with schizophrenia and nicotine use received single sessions of TMS applied to an individual-specific map of the left lateral parietal node of the DMN. Individuals first completed a Precision Targeting Visit to enable personalized TMS target selection. Participants then underwent three single sessions of TMS (intermittent theta burst stimulation, iTBS; continuous theta burst stimulation, cTBS; and sham) in a randomized, crossover design. At each TMS visit, participants reported their craving using 0-10 visual analog scale (VAS) and underwent resting-state fMRI immediately before and after receiving TMS. In the Comparative Effectiveness TMS study (Figure 1B), 62 individuals who use nicotine participated in this randomized, crossover TMS study comparing DMN-targeted TMS to left dorsolateral prefrontal cortex (L DLPFC)-targeted TMS. Participants were enrolled in two groups: schizophrenia (n=30) and non-psychosis control (n=32). Participants received 5 daily sessions of 1) DMN-Targeted cTBS and 2) L DLPFC-Targeted iTBS with pre-/post-TMS neuroimaging and VAS craving in a randomized, crossover design. Created with BioRender.com.

Surprisingly, it remains unknown if brain-behavior relationships observed with single TMS sessions persist following multiple TMS sessions. To test if *multiple* sessions would lead to an enduring effect on craving *and* if craving change would be related to this same pattern of bilateral parietal DMN connectivity change, we scaled up the TMS intervention to five sessions. Rather than comparing to sham, in Comparative Effectiveness TMS, we compared DMN-targeted TMS to a well-established, effective TMS target for nicotine use in the general population: the left dorsolateral prefrontal cortex (DLPFC) (11, 12) using a crossover design (Figure 1B). To compare the effectiveness of these targets, we tested these interventions in two groups of people using nicotine: 1) schizophrenia; and 2) a control group of individuals without psychosis. We hypothesized that DMN-targeted TMS would reduce craving more than DLPFC-targeted TMS for people with schizophrenia and that craving change would be associated with bilateral parietal DMN connectivity change.

It is presumed that TMS exerts its effects by inducing synaptic plasticity (13). Schizophrenia is recognized as both a disorder of impaired synaptic plasticity and of accelerated aging (14), which may affect response to TMS. In healthy participants, TMS has differential effects in older compared to younger individuals (15–18). Therefore, we tested if age would interact with diagnosis to affect response to TMS.

Our findings provide foundational support for bilateral parietal DMN connectivity as a novel mechanism by which TMS reduces craving in schizophrenia.

## Methods

### Protocol Optimization TMS in Schizophrenia

*Participants:* Fifteen individuals with schizophrenia aged 18-65 who use nicotine (confirmed for cigarette smokers by expired carbon monoxide ≥ 5ppm) were enrolled in this randomized, sham-controlled, crossover TMS-fMRI study (Table 1, Figure 1A, Supplemental Figure 1, NCT07190352). Participants provided written informed consent. Details in supplement.

**Table 1.** Demographics.

|  | **Protocol**<br>**Optimization TMS** | **Comparative Effectiveness TMS** |  |  |
| --- | --- | --- | --- | --- |
|  | **SZ + Nicotine**<br>**(n=11)** | **SZ + Nicotine**<br>**(n=30)** | **Non-Psychosis**<br>**Control + Nicotine**<br>**(n=32)** | **p** |
| **Age, y (SD)** | 38.73 (13.70) | 33.03 (11.46) | 37.13 (13.01) | .065 |
| **Sex, n (% Female)** | 8M/3F (27.3) | 5 (16.7) | 11 (35.5) | .041 |
| **Race, n (%)** |  |  |  |  |
| Black/African American | 2/11 (18.2) | 12 (40.0) | 1 (3.1) | <.0001 |
| Native American | 1/11 (9.1) | 0 (0.0) | 0 (0.0) |  |
| White | 8/11 (72.3) | 15 (50.0) | 28 (87.5) |  |
| Native Hawaiian or<br>Other Pacific Islander | 0/11 (0.0) | 1 (3.3) | 0 (0.0) |  |
| Asian | 0/11 (0.0) | 0 (0.0) | 0 (0.0) |  |
| Multiracial | 0/11 (0.0) | 2 (6.7) | 2 (6.3) |  |
| Other, not specified | 0/11 (0.0) | 0 (0.0) | 2 (6.3) |  |
| **Psychiatric Symptoms** |  |  |  |  |
| PANSS Total (SD) | 55.27 (11.84) | - | - |  |
| BPRS Total (SD) | - | 40.6 (12.0) | 30.1 (5.1) | <.0001 |
| **Nicotine Use** |  |  |  |  |
| Cigarette/Vape/Non-<br>combustible/Multiple, n | 8/1/0/2 | 12/10/1/7 | 13/13/3/3 | .41 |
| FTND* (SD) | 5.33 (1.32) | 4.63 (1.98) | 2.17 (1.98) | <.001 |
| Cigarettes/Day (SD) | 14.89 (6.67) | 9.57 (6.37) | 8.5 (7.17) | .69 |
| Expired CO* (SD) | 17.2 (9.0) | 16.53 (11.95) | 15.73 (17.72) | .84 |
| PSECDI (SD) | - | 12.14 (4.69) | 9.73 (5.22) | .24 |
| mL Vape Liquid (SD) | 2.35 (2.62) | - | - |  |
| Times Vaping/Day (SD) | - | 18.06 (20.54) | 11.32 (12.06) | .19 |
| **TMS Parameters** |  |  |  |  |
| AMT (SD) | 40.9 (6.82) | 44.4 (17.44) | 40.56 (8.70) | .28 |
| cTBS Intensity | 80% AMT | 100% AMT | 100% AMT | - |
| iTBS Intensity | 100% AMT | 100% AMT | 100% AMT | - |

AMT: Active Motor Threshold; CO: Carbon Monoxide; FTND: Fagerstrom Test for Nicotine Dependence; NMR: Nicotine Metabolite Ratio; PANSS: Positive and Negative Syndrome Scale; PSECDI: Penn State Electronic Cigarette Dependence Index. \*FTND and Expired CO reported only for combustible tobacco users. Vaping reported in times/day, defined as 15 puffs or 10 minutes as per PSECDI.

*Nicotine Assessments:* Nicotine use was assessed at each of five visits. Craving was assessed using 0-10 visual analogue scale (VAS) immediately before and after each TMS session.

*MRI Acquisition & Processing:* Imaging data were collected on a Siemens 3.0-T MRI system (Munich, Germany). Participants each completed 7 MRIs: baseline and immediately before and after each TMS session (Figure 1A). A total of 270 min of imaging data was collected per participant, including 140 min of resting-state imaging. Briefly, 1-mm^3^ T1-weighted anatomical scans and multiple 10-minute functional runs were acquired (TR 650ms, TE 34.80ms, flip angle 50 degrees, multiband acceleration factor 8, field of view = 207mm, 64 slices, 3-mm^3^ voxels, anterior to posterior phase-encoded). All resting-state scans went through a quality assurance procedure that included calculating framewise displacement (FD) and temporal signal to noise ratio (tSNR). Scans with mean FD>0.5 or tSNR<5th percentile of the sample distribution were excluded. Details in supplement.

*Protocol Optimization TMS Protocol:* Individuals received single sessions of theta-burst stimulation (Figure 1A & Supplemental Figure 1) applied to an individualized left parietal DMN target (see *Individualized DMN Target* in Supplement and Supplemental Figure 2) (10). Individuals received one session of iTBS (600 pulses, 100% Active Motor Threshold, AMT), cTBS (600 pulses, 80% AMT as per (19)), and sham (coil flipped 180 degrees using 100% AMT iTBS protocol, 600 pulses). Order was randomized, and participants were blinded. Details in supplement.

*Longitudinal Neuroimaging Analysis:* We used the SPM Sandwich Estimator Toolbox for Longitudinal & Repeated Measures Data (SwE, https://www.nisox.org/Software/SwE/) to identify patterns of connectivity change from the TMS target. We generated connectivity maps from the left lateral parietal DMN region (using seed from (20)) for each MRI then created difference maps for each TMS session by subtracting post-TMS map - pre-TMS map. We used the SwE toolbox to model the effect of craving change on connectivity change using 20,000 bootstraps then performed voxel-wise tests with FDR correction. Mean connectivity values were extracted from individuals from the largest cluster identified at the group level and used to visualize results.

***Comparative Effectiveness TMS in Schizophrenia and Non-Psychosis Controls*** *Participants:* Ninety individuals aged 18-65 who use nicotine were enrolled. Participants were recruited in two groups: 1) Schizophrenia and 2) Non-psychosis control (Table 1, Figure 1B, Supplemental Figure 3, NCT06389266). Individuals in the schizophrenia group had a DSM-V diagnosis of schizophrenia or schizoaffective disorder, while individuals in the non-psychosis control group had no lifetime history of psychosis. Participants provided written informed consent. Details in supplement.

*Nicotine Assessments:* Nicotine use was assessed at each visit. Craving was assessed using 0-10 VAS at each MRI visit.

*MRI Acquisition & Processing:* Imaging data were collected on 3.0-T Philips Intera Achieva MRI scanner (Philips Healthcare, Andover, MA). Participants completed MRIs the week before and after TMS. Post-TMS scans were scheduled within 8 days after TMS (mean 4.39 (SD 1.61) days). Briefly, 1-mm^3^ T1-weighted anatomical scans and multiple 10-minute functional runs were acquired (TR 2000ms, TE 28.0ms, flip angle 90 degrees, field of view = 240mm, 38 slices, 3-mm^3^ voxels, anterior to posterior phase-encoded). Scans were processed using the same procedure described above. Details in supplement.

*Comparative Effectiveness TMS Protocol:* In this randomized, crossover trial, individuals received five daily sessions of 1) DMN-Targeted cTBS and 2) left DLPFC-Targeted iTBS with pre-/post-TMS neuroimaging. TMS order was randomized, and interventions were separated by a two-week washout period. DMN-Targeted cTBS (600 pulses, 100% AMT) was applied to an individualized left parietal DMN target (see *Individualized DMN Target* in Supplement). Left DLPFC-Targeted iTBS (600 pulses, 100% AMT) was anatomically targeted using MNI coordinates for the average 5cm rule (x = −41, y = +16, z = +54) (21). Details in supplement.

*Region-of-Interest (ROI) Neuroimaging Analysis:* For Comparative Effectiveness TMS, we calculated connectivity from each TMS target to our previously identified right parietal DMN region. To do this, we extracted time courses from 6mm spheres placed at 1) our left lateral parietal DMN TMS target (x = −46, y = −66, z = +30) (20) and 2) our left DLPFC TMS target (x = −41, y = +16, z = +54) to our previously identified right parietal DMN cluster (Figure 2A). The BOLD signal time courses from ROIs were correlated with each other and z-transformed to generate ROI-to-ROI connectivity values for analysis.

**Figure 2.**
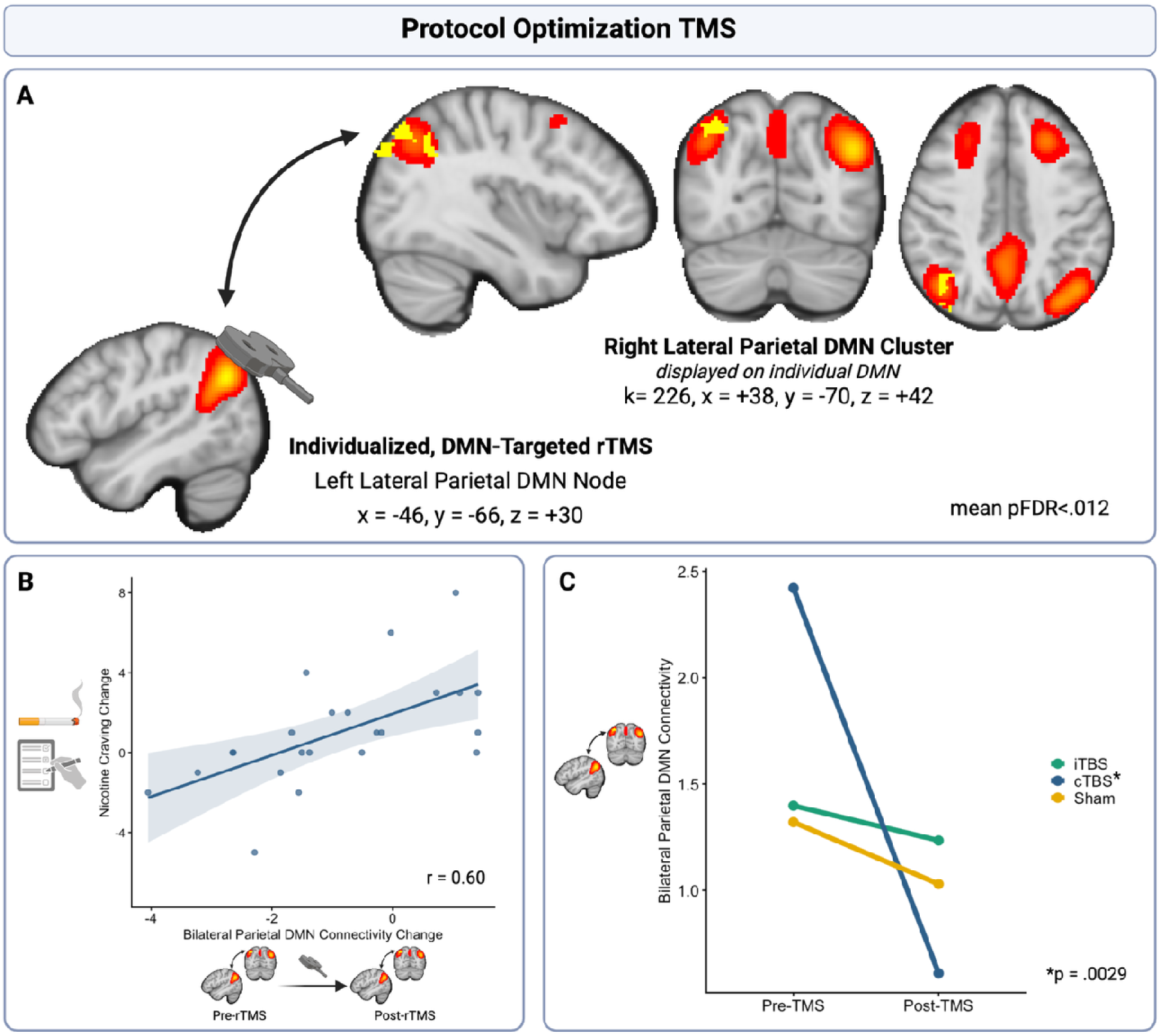
Craving Change is Related to Bilateral Parietal DMN Connectivity Change in Protocol Optimization TMS. In our Protocol Optimization TMS study, craving change was related to connectivity change within the DMN. The largest cluster was located in the right parietal cortex (Figure 2A, cluster k = 226, located at MNI x = +38, y = −70, z = +42, mean pFDR<0.012). As connectivity between the left lateral parietal DMN and right parietal cluster increased, craving also increased (r = 0.60, Figure 2B). There was a significant treatment by tim interaction (p=.0012) and main effect of treatment (p<0.001) on bilateral parietal connectivity. Adjusted tests of treatment effect showed cTBS reduced bilateral parietal connectivity (p_adj_=.0029; Figure 2C). iTBS and sham did not significantly affect connectivity (p_adj_>0.05).

### Statistical Analysis

Chi-square or Fisher’s Exact tests (if cell counts<5) were used to compare categorical variables and Welch T-tests were used to compare continuous outcomes between groups (Table 1).

For the Protocol Optimization TMS neuroimaging analysis, mean connectivity values extracted from the largest cluster from voxel-wise analyses were modeled using linear mixed effects models, including the interaction between time and stimulation condition (iTBS, cTBS, sham) and a random intercept for subject. All mixed models tests were performed using Type 2 sum of squares and Satterthwaite’s adjustment for degrees of freedom. A linear mixed effects model with robust standard errors was used to model connectivity by treatment time (pre- and post-stimulation), TMS condition (iTBS, cTBS, sham) and their interaction, including a random effect for subject. Tests of treatment effect for each stimulation condition were adjusted across groups using Tukey correction. To investigate the effect of age on the association between craving change (post-pre) and connectivity change (post-pre), we used a linear mixed effects model to fit craving change (post-pre) by change in connectivity, the connectivity change*age interaction and a random intercept for subject.

In Comparative Effectiveness TMS, we investigated the association of TMS target with craving change by using linear mixed effects models to fit craving change (post-pre) by pre-TMS craving and TMS target. To test for group effects on the association between craving change and TMS target, we used linear mixed effects models to predict craving change by pre-TMS craving, TMS target, group, and the TMS target*group interaction. We investigated the association of TMS target with connectivity change by using linear mixed effects models to fit connectivity change by pre-TMS connectivity, TMS target, and age. To test for group effects on the association between connectivity change and TMS target, we used linear mixed effects models to predict connectivity change by pre-TMS connectivity, TMS target, group, age, and the TMS target*group interaction.

To investigate how associations between craving change (post-pre) and connectivity change (post-pre) differ by age and group, we used linear mixed effects models to fit craving change (post-pre) by change in connectivity controlling for pre-TMS connectivity and craving, group, age, and their 3-way interaction including a random intercept for subject. All analyses were conducted in RStudio (Version 2023.03.1+446).

## Results

### Protocol Optimization TMS in Schizophrenia

Ten participants with schizophrenia and daily nicotine use completed our randomized, controlled crossover study of individualized, left parietal DMN-targeted TMS (Figure 1A). Effects on craving were previously reported (10). Details in supplement.

### Craving Change is Related to Bilateral Parietal Connectivity Change

We performed voxel-wise analyses to identify regions where left lateral parietal DMN connectivity changed with craving, agnostic of TMS intervention. Craving change was related to connectivity change within the DMN. The largest cluster was located in the right parietal cortex (cluster k = 226, located at MNI x = +38, y = −70, z = +42, mean pFDR<0.012, Figure 2A, Supplemental Table 1). As bilateral parietal connectivity increased, craving also increased (r = 0.60, Figure 2B).

There was a significant treatment by time interaction (X^2^=13.39, df=2, p=.0012) and main effect of treatment (X^2^=15.68, df=2, p<.001) on bilateral parietal connectivity. Tukey-adjusted tests of treatment effect showed cTBS significantly reduced bilateral parietal connectivity (Estimate= - 1.80, t=3.18, df=37.4, p_adj_=.0029; Figure 2C) with insufficient evidence for iTBS (Estimate= - 0.28, t=0.69, df=37.6, p_adj_=.49) and sham (Estimate= −0.17, t=0.32, df=38.0, p_adj_=.75). There was a nonsignificant main effect of time (X^2^=0.92, df=1, p=.34). In a mixed effects model predicting craving change, there was no interaction between bilateral parietal DMN connectivity change and age (p=0.14, Supplemental Table 2).

### Comparative Effectiveness TMS in Schizophrenia and Non-Psychosis Controls

Sixty-two participants using nicotine (30 schizophrenia, 32 non-psychosis control) completed our randomized, crossover study of DMN-targeted TMS compared to left DLPFC-targeted TMS (Figure 1B, Supplemental Figure 3). Details in supplement.

### TMS Reduces Nicotine Craving

We used mixed effects models to study the association between craving change and TMS target (Supplemental Table 3). There was no significant main effect of TMS target (beta=0.11, standard error (SE)=0.32, T(60.38)=0.34, p=0.73) suggesting craving change was similar between the two treatments. The treatment led to significant reductions in craving for both TMS targets (DLPFC: est=-0.81, SE=0.25, 95% CI=[−1.30, −0.32], T(117.61)=-3.26, p=0.0015; DMN: est=-0.70, SE=0.25, 95% CI=[-1.19, −0.21], T(117.38)=-2.83, p=0.0054, Supplemental Table 4).

We also used mixed effects models to determine whether the association between craving change and TMS target differed by diagnostic group (Supplemental Table 5). There was no significant interaction of TMS target with diagnosis (beta=0.13, SE=0.65, T(59.61)=0.20, p=0.84, Figure 3B) suggesting the effect of TMS target was similar across the diagnostic groups.

**Figure 3.**
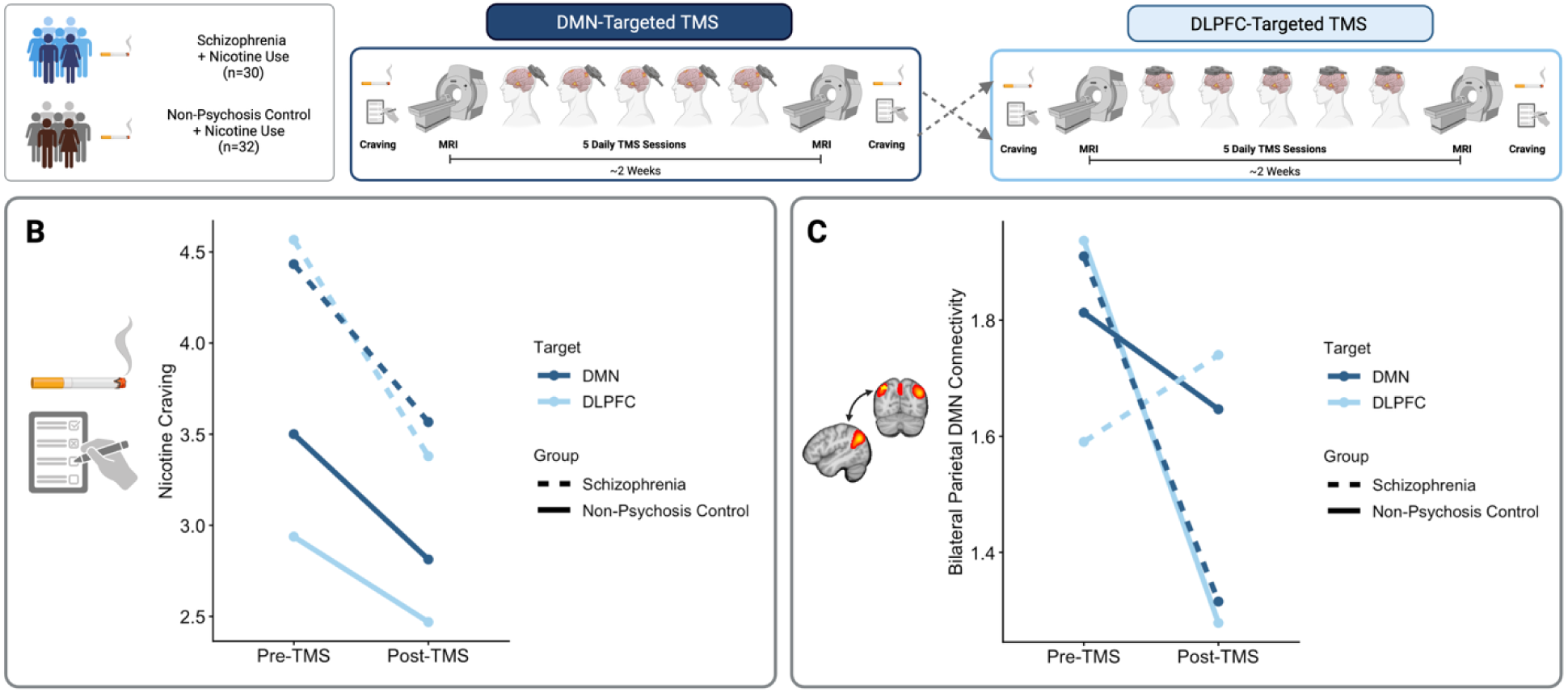
In Comparative Effectiveness TMS, TMS Reduces Nicotine Craving and Bilateral Parietal DMN Connectivity. In our Comparative Effectiveness TMS study comparing five sessions of DMN-targeted and L DLPFC-targeted TMS in nicotine using people with schizophrenia and a non-psychosis control group, both TMS targets reduced nicotine craving (DLPFC: p=0.0015; DMN: p=0.0054). There was no significant interaction of TMS target with diagnosis (p=0.84, Figure 3B) suggesting the effect of TMS target was similar across the diagnostic groups. Both TMS interventions reduced bilateral parietal DMN connectivity (DLPFC: p=0.024; DMN: p=0.023). There was a significant interaction of TMS target with diagnosis (p=0.016, Figure 3C). TMS led to significant reductions in bilateral parietal DMN connectivity when applied to the DLPFC target for the non-psychosis control group (p=0.0027) and when applied to the DMN target in the schizophrenia group (p=0.0089). There were no changes in connectivity with TMS applied to the DLPFC in the schizophrenia group or the DMN target in the control group (p>0.05).

### TMS Reduces Bilateral Parietal DMN Connectivity

We used mixed effects models to test the association between change in bilateral parietal DMN connectivity with TMS target and age (Supplemental Table 6). There was no significant main effect of TMS target (beta=0.00, SE=0.20, T(50.11)=0.00, p=1.0) or age (beta=-0.00, SE=0.01, T(46.91)=-0.27, p=0.79), suggesting the connectivity change was similar between the two treatments and not affected by age. The treatment led to significant reductions in bilateral parietal DMN connectivity for both TMS targets (DLPFC: est= −0.33, SE=0.15, 95% CI=[−0.62, −0.044], T(95.98)=-2.29, p=0.024; DMN: est=-0.33, SE=0.14, 95% CI=[−0.61, −0.050], T(95.98)=-2.34, p=0.022, Supplemental Table 7). Pre-TMS bilateral parietal DMN connectivity did not differ by group or age (Supplemental Figure 4).

We used mixed effects models to determine whether the association between bilateral parietal DMN connectivity change with TMS target differed by diagnostic group (Supplemental Table 8). There was a significant interaction of TMS target with diagnosis (beta=-1.00, SE=0.40, T(50.38)=-2.49, p=0.016, Figure 3C). TMS led to significant reductions in bilateral parietal DMN connectivity when applied to the DLPFC target for the non-psychosis control group (est=-0.58, SE=0.19, 95% CI=[−0.96, −0.21], T(93.90)=-3.08, p=0.0027) and when applied to the DMN target in the schizophrenia group (est=-0.56, SE=0.21, 95% CI=[−0.98, −0.14], T(93.86)=-2.67, p=0.0089). There were no connectivity changes with TMS applied to the DLPFC in the schizophrenia group (est=0.0047, SE=0.22, 95% CI=[-0.43, 0.44], T(93.83)=0.022, p=0.98) or the DMN target in the control group (est=-0.15, SE=0.19, 95% CI=[-0.52, 0.23], T(93.90)=-0.77, p=.44, Supplemental Table 9).

To test the specificity of the bilateral parietal DMN connectivity change, we tested if connectivity between the DLPFC TMS target and right parietal DMN region changed with TMS. In separate mixed effects models predicting DLPFC-right parietal DMN connectivity change, there was no effect of TMS target (beta=0.25, SE=0.19, T(49.14)=1.32, p=0.19) or interaction between TMS target and diagnosis (beta=-0.40, SE=0.39, T(50.55)=-1.03, p=0.31, Supplemental Tables 10 & 11).

### Craving Change is Related to Bilateral Parietal Connectivity Change

We used mixed effects models to test if craving change was associated with bilateral parietal DMN connectivity change, group, and age (Supplemental Table 12). There was a significant interaction of connectivity change, group, and age (beta=0.12, SE=0.04, T(74.94)=3.33, p=0.001, Figure 4). To understand this interaction, we estimated slopes of bilateral parietal DMN connectivity change predicting craving change for each group at the 25^th^ and 75^th^ percentiles of age (24 and 46) from our mixed effects model. In schizophrenia, increased connectivity was associated with increased craving in older individuals (est=2.32, SE=0.69, 95% CI=[0.95, 3.70], T(84.6)=3.37, p=.0011), while in younger individuals with schizophrenia, increased connectivity was associated with decreased craving at a trend level (est=-0.44, SE=0.25, 95% CI=[-0.94, 0.063], T(79.5)=-1.74, p=.086). In the control group, connectivity change was not associated with craving change at any age (Supplemental Table 13).

**Figure 4.**
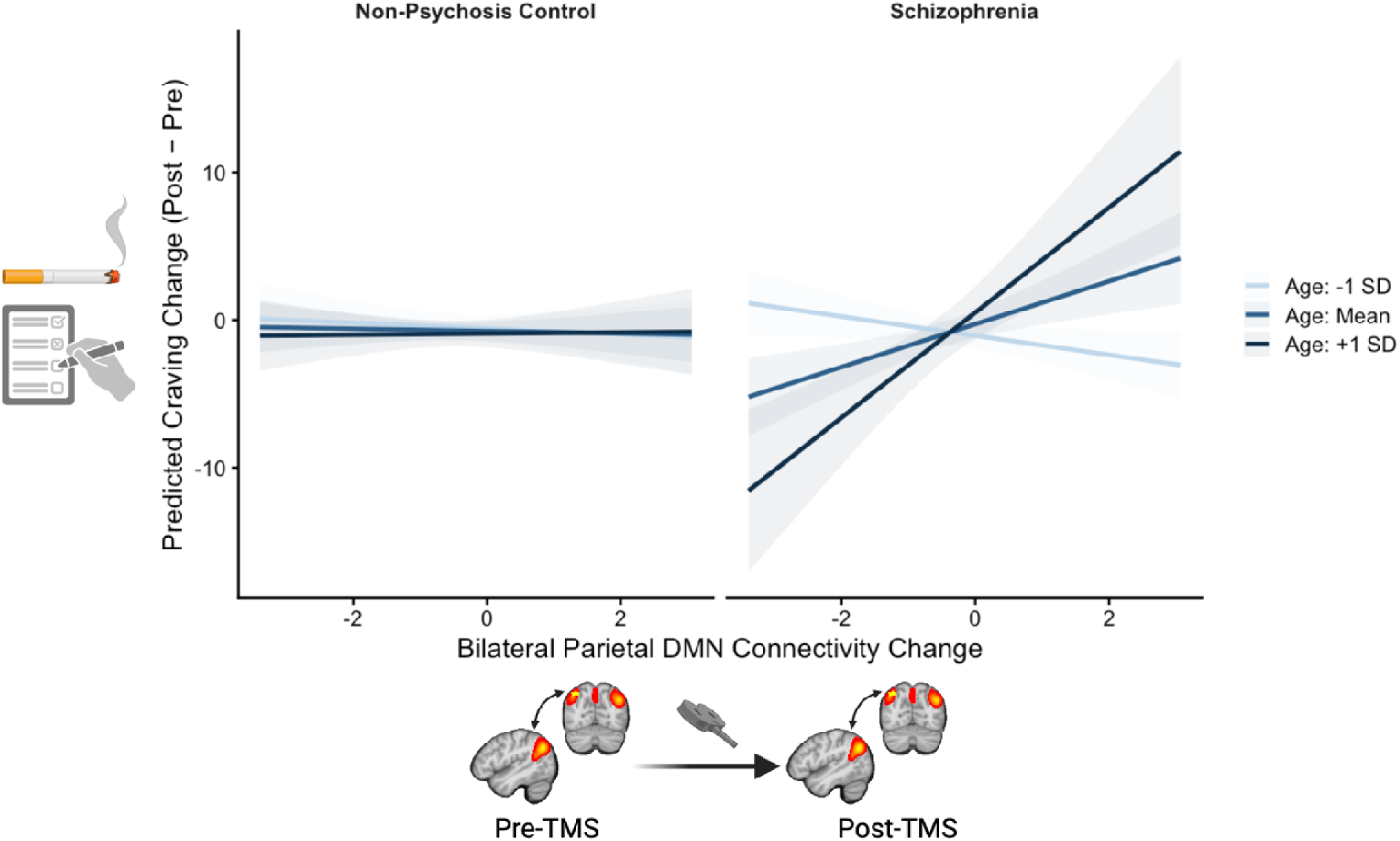
Craving Change is Related to Bilateral Parietal DMN Connectivity Change, Group, and Age in Comparative Effectiveness TMS. In our Comparative Effectiveness TMS study comparing five sessions of DMN-targeted and L DLPFC-targeted TMS in nicotine using people with schizophrenia and a non-psychosis control group, there was a significant interaction of bilateral parietal connectivity change, group, and age (p=0.001) on craving change. To understand this interaction, we estimated simple slopes (marginal trends) of bilateral parietal DMN connectivity change predicting craving change from our mixed effects model. Slopes were computed separately by group at the 25^th^ and 75^th^ percentiles of age (24 and 46 years). In schizophrenia, increased connectivity was significantly associated with increased craving in older individuals (p=0.001), while in younger individuals, increased connectivity was associated with decreased craving at a trend level (p=0.086). In the control group, connectivity change was not associated with craving change at any age.

To test the specificity of the association between craving change and bilateral parietal DMN connectivity change, we used mixed effects models to test if craving change was associated with L DLPFC-right parietal DMN connectivity change. There was no significant interaction of DLPFC-right parietal connectivity change, group, and age (beta=-0.04, SE=0.03, T(79.30)=-1.20, p=0.23, Supplemental Table 14).

## Discussion

Tobacco use is a public health crisis for people with schizophrenia, but the field has been unable to elucidate the underlying pathology or to develop effective smoking cessation treatments for this vulnerable population. We used two neuromodulation experiments to identify a novel brain mechanism for nicotine craving that is unique to schizophrenia, persists from single to multiple TMS sessions, and provides mechanism for a schizophrenia-specific smoking cessation treatment.

In Protocol Optimization TMS, we used single sessions of DMN-targeted TMS with pre-post neuroimaging to show that craving change is related to bilateral parietal DMN connectivity change and that DMN-targeted cTBS reduces bilateral parietal DMN connectivity.

To test if this craving-connectivity relationship persisted after multiple sessions of TMS, we scaled up the intervention from single to multiple TMS sessions with pre-post neuroimaging in Comparative Effectiveness TMS. We compared effects of DMN-targeted cTBS and left DLPFC-targeted iTBS in nicotine-using people with schizophrenia and non-psychosis controls. Both interventions reduced craving and bilateral parietal DMN connectivity. We then observed that craving change was related to the interaction between bilateral parietal DMN connectivity change, group, and age, such that the relationship between craving change and bilateral parietal DMN connectivity change was specific to schizophrenia and varied by age. In older people with schizophrenia, increased bilateral parietal DMN connectivity was associated with *increased* craving. However, in younger people with schizophrenia, increased bilateral parietal DMN connectivity was associated with *reduced* craving. At baseline, bilateral parietal DMN connectivity was not associated with group or age, suggesting that this was in fact a difference in *response* to TMS, rather than baseline differences. Although this age effect has not been specifically observed in schizophrenia, it is consistent with differential responses to TMS based on age in healthy populations (15–18). In summary, we demonstrated that even after multiple TMS sessions and over multiple weeks, craving change was reliably related to bilateral parietal DMN connectivity change in schizophrenia.

So why the parietal DMN? There are several potential explanations. It is well-documented that TMS consistently generates neural changes in contralateral homologous brain regions, dating back to the infancy of TMS research. TMS applied to the motor cortex (M1) affects motor-evoked potentials and EEG signals and increases PET activity observed in contralateral M1 (22–26). Moreover, in healthy controls, TMS applied to the left inferior parietal lobule (IPL, i.e., our approximate DMN target) affects functional connectivity to the right IPL (27).

Although not commonly recognized as part of canonical reward circuitry, the parietal cortex is frequently implicated in craving. The posterior parietal cortex activates following smoking cue exposure (28–33). Among treatment-seeking smokers, cue-elicited responses in parietal cortex were associated with relapse vulnerability (34), and cue reactivity patterns in parietal cortex are linked to individual differences in nicotine dependence and relapse risk (35). Together, this literature shows that craving involves attention and imagery systems governed by parietal cortex. Therefore, it is possible that the reductions in bilateral parietal DMN connectivity we observed in our TMS experiments are indicative of decreased attention paid to nicotine craving. The associations we observed between craving change and bilateral parietal DMN connectivity change are consistent with prior work that links craving to within-DMN connectivity (36) and that acute nicotine administration (which reduces craving) reduces within-DMN connectivity (6, 37, 38).

This series of experiments has several major strengths. This combination of studies demonstrating a common therapeutic mechanism represents a major advancement in the therapeutic development pipeline. The Protocol Optimization TMS experiment uses a sophisticated 3-arm crossover design with precision functional imaging of 70 scans in symptomatic individuals with schizophrenia to test mechanism and select an optimal TMS protocol. The Comparative Effectiveness TMS experiment then scales up this optimal TMS intervention to 5 sessions to test therapeutic effect and validate its mechanism over a multi-week timescale. A unique strength of this study is its use of a gold-standard active comparison TMS intervention. Left DLPFC-targeted TMS has well-established therapeutic effects on nicotine craving, so it serves as a more rigorous and clinically-relevant comparator than sham. Finally, the use of two groups of nicotine users allows for comparison of therapeutic responses by diagnosis. The use of a non-psychosis control group (i.e., individuals with other psychiatric diagnoses) provides a real-world comparison arm, as the majority of individuals who use nicotine have a co-occurring psychiatric disorder (39, 40).

Our studies have several limitations as well. The Protocol Optimization TMS study was relatively small, though did include a high volume of within-subject data, and did not include a non-psychosis control group. Although the inclusion of an effective, active control intervention was a strength in the Comparative Effectiveness TMS study, there was no sham intervention. Future studies should scale up the sessions of DMN-targeted cTBS in a large, sham-controlled trial for smoking cessation.

In conclusion, these findings support a novel, schizophrenia-specific brain mechanism underlying nicotine craving that can be targeted for therapeutic benefit. This represents a substantial advancement in the therapeutic development pipeline, which often lacks consistent mechanistic evidence. These findings provide hope for development of effective smoking cessation treatments for people with schizophrenia.

## Data Availability

All data produced in the present study may be available upon reasonable request.

## Acknowledgments

The authors would like to thank the participants in these studies.

