## Supplement for "A Novel Therapeutic Mechanism for Nicotine Craving in Schizophrenia"

**Supplemental Material**

**Supplemental Methods**

***Protocol Optimization TMS in Schizophrenia***

*Participants:* Fifteen individuals with schizophrenia or schizoaffective disorder aged 18-65 who use nicotine (confirmed for cigarette smokers by expired carbon monoxide ≥ 5ppm) were enrolled in this randomized, sham-controlled, crossover TMS-fMRI study. Diagnosis was confirmed by DSM-V SCID interview (1) and clinical information obtained from outpatient psychiatric providers. For one month prior to enrollment, individuals received outpatient care, with no hospitalizations or changes to their psychiatric medication regimens. Individuals were excluded if they had DSM-V intellectual disability, substance use disorder (other than nicotine) in the past 3 months, a progressive or genetic neurologic disorder, history of significant head trauma, history of seizures or neurosurgical procedures, implanted devices, gross organic pathology on neuroimaging, contraindications to MRI or TMS, or current pregnancy. All participants provided written informed consent in accordance with the Beth Israel Deaconess Medical Center Institutional Review Board.

*Nicotine Assessments*: Nicotine dependence was assessed using the Fagerstrom Test for Nicotine Dependence (FTND) (2) and Penn State Electronic Cigarette Dependence Index (PSECDI) (3).

*MRI Data Processing:* Anatomical images were segmented into gray matter, white matter and cerebrospinal fluid (CSF) with the Computational Anatomy Toolbox 12 (CAT12, version 12.5; [http://www.neuro.uni-jena.de/cat/](https://nam12.safelinks.protection.outlook.com/?url=http%3A%2F%2Fwww.neuro.uni-jena.de%2Fcat%2F&data=05%7C01%7Cheather.b.ward%40vumc.org%7C53146c81d5534edf7b5208db2189eebc%7Cef57503014244ed8b83c12c533d879ab%7C0%7C0%7C638140648518151829%7CUnknown%7CTWFpbGZsb3d8eyJWIjoiMC4wLjAwMDAiLCJQIjoiV2luMzIiLCJBTiI6Ik1haWwiLCJXVCI6Mn0%3D%7C3000%7C%7C%7C&sdata=Fsce8FRawoYi0jL0FjzH%2BUsdG8P5cAj4Wzyuk79pHuo%3D&reserved=0)). Resting-state scans were preprocessed in SPM12 and Matlab and were (1) realigned to a mean scan, (2) coregistered with the native space structural scan, then (3) underwent simultaneous resting-state denoising procedures via regression for removal of: sine/cosine terms for a bandpass filter 0.01–0.1 Hz; the mean GM signal; the first 6 principal components of the combined WM/CSF compartment; and the 6 estimated translation and rotation head movement parameters plus their 6 first derivatives. All resting state scans went through a quality assurance procedure that included calculating framewise displacement (FD) and temporal signal to noise ratio (tSNR). Scans with a mean FD > 0.5 or a tSNR lower than the 5th percentile of the distribution of the entire sample were excluded from further analysis. After quality control, there were pre- and post-TMS scans for 8 cTBS, 9 iTBS, and 6 sham sessions across 9 subjects.

*TMS Protocol Motor Threshold Determination:* Participants had motor threshold determination at their first TMS visit. Single pulse and repetitive stimulation was performed with a MagPro stimulator (MagVenture) equipped with a biphasic figure-of-eight coil. To obtain an active motor threshold, single pulses were used in the following manner: Electromyographic activity (EMG) was recorded using surface electrodes attached to the skin to measure motor evoked potentials (MEP) during the motor threshold assessment. The TMS coil was placed on the scalp. Single TMS pulses were applied over the hand area of the left motor cortex and individually localized for each participant based on the optimal position for eliciting a motor evoked potential. Neuronavigation (Brainsight, Rogue Research, Inc.) was used to record the motor ‘hot-spot.’ Resting and active motor threshold (RMT; AMT) were obtained by following recommendations from the International Federation of Clinical Neurophysiology.

*Protocol Optimization TMS Protocol:* Individuals received single sessions of theta-burst stimulation (Figure 1A & Supplemental Figure 1) applied to an individualized left parietal DMN target (see *Individualized DMN Target* below and Supplemental Figure 2) with neuroimaging collected immediately before and after each session (4). Individuals received one session of iTBS (600 pulses, 100% Active Motor Threshold, AMT), cTBS (600 pulses, 80% AMT as per (5)), and sham (coil flipped 180 degrees using 100% AMT iTBS protocol, 600 pulses) on separate days, separated by at least 2 days to avoid carryover effect (median 6.5 days, mean 13.8 days (SD 22.2), range 2-96 days). TMS was applied using a MagPro X100 stimulator and active figure-of-8 coil (Cool B65, MagVenture, Denmark) held tangentially to the scalp with the handle at 45 degrees. TMS was applied in the standard theta-burst pattern (3 pulses at 50-Hz repeated at a rate of 5-Hz) (6). Order was randomized, and participants were blinded.

***Comparative Effectiveness TMS***

*Participants:* Ninety individuals aged 18-65 who use nicotine were enrolled. Participants were recruited in two groups: 1) Schizophrenia and 2) Non-psychosis control (Figure 1B, NCTNCT06389266). Individuals in the schizophrenia group had a diagnosis of schizophrenia or schizoaffective disorder (n=31) confirmed by DSM-V SCID interview (1) and clinical information obtained from outpatient psychiatric providers. Individuals in the non-psychosis control group had no lifetime history of psychosis (n=32) confirmed by DSM-V SCID Part B interview (1). For one month prior to enrollment, individuals received outpatient care, with no hospitalizations or changes to their psychiatric medication regimens. Individuals were excluded if they had DSM-V intellectual disability, substance use disorder (other than nicotine) in the past 3 months confirmed by DSM-V SCID Part E interview, a progressive or genetic neurologic disorder, history of significant head trauma, history of seizures or neurosurgical procedures, implanted devices, gross organic pathology on neuroimaging, contraindications to MRI or TMS, or current pregnancy. All participants provided written informed consent in accordance with the Vanderbilt University Medical Center Institutional Review Board.

*Nicotine Assessments*: Nicotine dependence was assessed using the FTND (2) and PSECDI (3).

*MRI Acquisition and Data Processing:* Imaging data were collected on 3.0-T Philips Intera Achieva MRI scanner (Philips Healthcare, Andover, MA). Participants completed MRI scans the week before and after the TMS intervention. As the durability of 5 TMS sessions is unknown, post-TMS scans were scheduled within 8 days after TMS (mean 4.39 (SD 1.61) days). Briefly, 1-mm^3^ T1-weighted anatomical scans and multiple 10-minute functional runs were acquired (TR 2000ms, TE 28.0ms, flip angle 90 degrees, field of view = 240mm, 38 slices, 3-mm^3^ voxels, anterior to posterior phase-encoded). Scans were processed according to the same procedure described above for the Protocol Optimization TMS study: Anatomical images were segmented into GM, WM, and CSF with the Computational Anatomy Toolbox 12 (CAT12, version 12.5; [http://www.neuro.uni-jena.de/cat/](https://nam12.safelinks.protection.outlook.com/?url=http%3A%2F%2Fwww.neuro.uni-jena.de%2Fcat%2F&data=05%7C01%7Cheather.b.ward%40vumc.org%7C53146c81d5534edf7b5208db2189eebc%7Cef57503014244ed8b83c12c533d879ab%7C0%7C0%7C638140648518151829%7CUnknown%7CTWFpbGZsb3d8eyJWIjoiMC4wLjAwMDAiLCJQIjoiV2luMzIiLCJBTiI6Ik1haWwiLCJXVCI6Mn0%3D%7C3000%7C%7C%7C&sdata=Fsce8FRawoYi0jL0FjzH%2BUsdG8P5cAj4Wzyuk79pHuo%3D&reserved=0)). Resting-state scans were preprocessed in SPM12 and Matlab and were (1) realigned to a mean scan, (2) coregistered with the native space structural scan, then (3) underwent simultaneous resting-state denoising procedures via regression for removal of: sine/cosine terms for a bandpass filter 0.01–0.1 Hz; the mean GM signal; the first 6 principal components of the combined WM/CSF compartment; and the 6 estimated translation and rotation head movement parameters plus their 6 first derivatives. All resting-state scans went through a quality assurance procedure that included calculating framewise displacement (FD) and temporal signal to noise ratio (tSNR). Scans with mean FD>0.5 or tSNR<5th percentile of the sample distribution were excluded. After quality control, there were 200 pairs of pre-/post-TMS scans for analysis.

*TMS Protocol Motor Threshold Determination:* Participants had motor threshold determination at their first TMS visit. Single pulse and repetitive stimulation was performed with a MagPro stimulator (MagVenture, Denmark) equipped with a biphasic figure-of-eight coil. To obtain an active motor threshold, single pulses were used in the following manner: Electromyographic activity (EMG) was recorded using surface electrodes attached to the skin to measure motor evoked potentials (MEP) during the motor threshold assessment. The TMS coil was placed on the scalp. Single TMS pulses were applied over the hand area of the left motor cortex and individually localized for each participant based on the optimal position for eliciting a motor evoked potential. Neuronavigation (Brainsight, Rogue Research, Inc.) was used to record the motor ‘hot-spot.’ Resting and active motor threshold (RMT; AMT) were obtained by following recommendations from the International Federation of Clinical Neurophysiology.

*Comparative Effectiveness TMS Protocol:* In this randomized, crossover trial, individuals received 5 daily sessions of 1) DMN-Targeted cTBS and 2) L DLPFC-Targeted iTBS with pre-/post-TMS neuroimaging. TMS order was randomized, and interventions were separated by a two-week washout period. DMN-Targeted cTBS (600 pulses, 100% AMT) was applied to an individualized left parietal DMN target (see *Individualized DMN Target* below). L DLPFC-Targeted iTBS (600 pulses, 100% AMT) was anatomically targeted using MNI coordinates for the average 5cm rule for scalp-based DLPFC targeting (x = -41, y = +16, z = +54) (7) with Brainsight neuronavigation software (Rogue Research, Inc.). TMS was applied using a MagPro X100 stimulator and an active figure-of-8 coil (Cool B65, MagVenture, Denmark) held tangentially to the scalp with the handle at 45 degrees. TMS was applied in the standard theta-burst pattern (3 pulses at 50-Hz repeated at a rate of 5-Hz) (6). Details in supplement.

***Individualized DMN Target***

To modulate the DMN, we selected a personalized target in the left lateral parietal region of the DMN. This target was selected based on prior work in healthy populations showing that applying TMS to this DMN node effectively modulated DMN connectivity (8). To identify an individualized DMN map for TMS targeting, a standard DMN template (9) was warped into native space and applied to the participant’s pre-TMS scan. In each participant, the resultant connectivity maps yielded a correlation cluster in the left posterior inferior parietal lobule (IPL, Supplemental Figure 2). A target was placed in the averaged center of the left posterior IPL correlation cluster (formed from the overlay of the left posterior IPL clusters derived from connectivity maps) on the cortical surface using Brainsight neuronavigation software (Rogue Research, Inc.). This individualized left parietal DMN region served as the TMS target for both the Protocol Optimization TMS and Comparative Effectiveness TMS studies.

**Supplemental Results**

***Protocol Optimization TMS in Schizophrenia***

*Single Sessions of DMN-Targeted TMS are Safe and Well-Tolerated in Schizophrenia*

As previously reported (4), TMS was safe and well-tolerated. All 10 participants who were randomized completed all three TMS/fMRI visits (Supplemental Figure). No serious adverse events were observed. Participants endorsed the following side effects: headache or neck pain (n=4) and trouble concentrating (n=5). One participant experienced hypomania that responded to medication adjustment. Four participants reported lower anxiety, and one participant reported improved mood.

***Comparative Effectiveness TMS in Schizophrenia and Non-Psychosis Controls***

*Multiple Sessions of DMN- and DLPFC-Targeted TMS are Safe and Well-Tolerated in Schizophrenia and Non-Psychosis Controls*

TMS was safe and well-tolerated. A total of 63 participants were randomized, and 62 completed the study (Supplemental Figure). One participant in the schizophrenia dropped out during TMS due to reported insomnia. No serious, unexpected, adverse events were observed.

*Pre-TMS Bilateral Parietal DMN Connectivity Does Not Differ by Group*

In a linear mixed effects model predicting pre-TMS bilateral parietal DMN connectivity neither age, group, nor their interaction were significant predictors (p>.05, Supplemental Figure 4 and Supplemental Table 15).

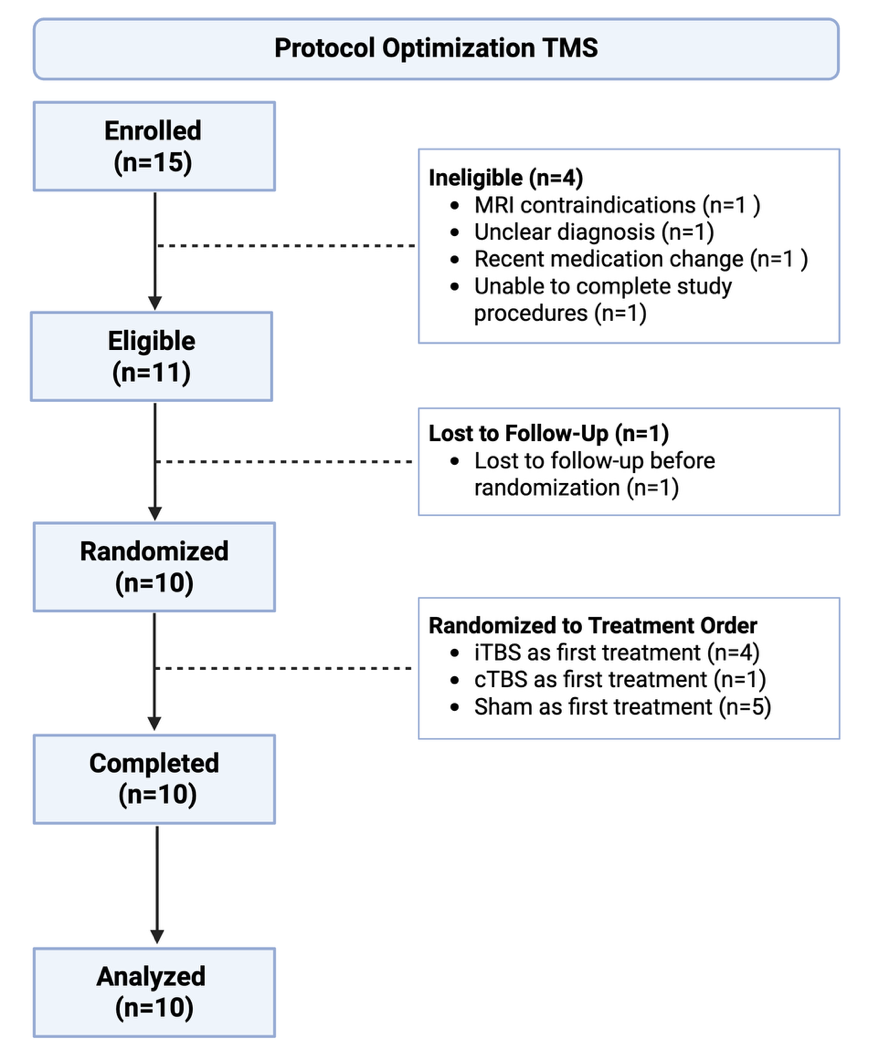
**Supplemental Figures and Tables:**

**Supplemental Figure 1. Protocol Optimization TMS Consort Diagram.** Fifteen participants with schizophrenia or schizoaffective disorder aged 18-65 who use nicotine were enrolled in this randomized, sham-controlled crossover study. Ten participants completed the study and provided data for analysis.

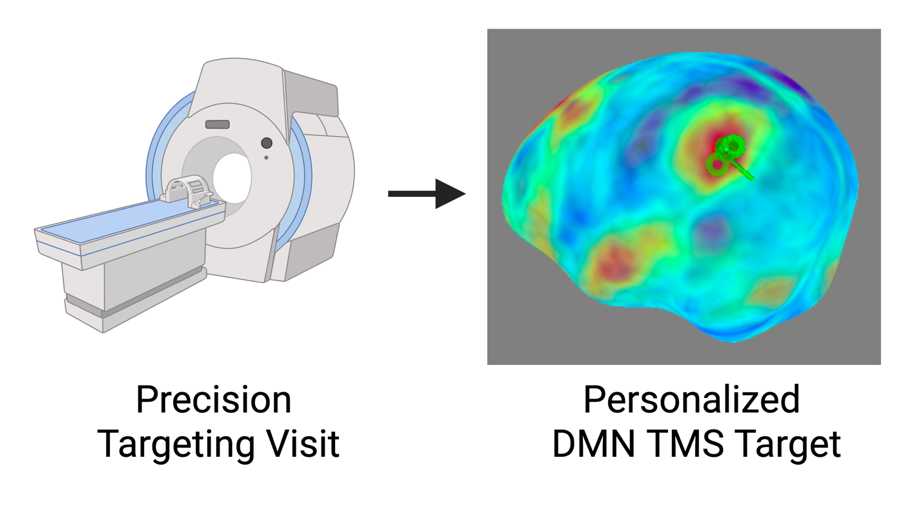

**Supplemental Figure 2. Individualized Default Mode Network TMS Target***:* The Default Mode Network (DMN) target was identified using the same methods for the Protocol Optimization TMS and Comparative Effectiveness TMS interventions. For our DMN target, we selected the left lateral parietal DMN, as it is a DMN region that is readily identifiable across all individuals and has been successfully used to modulate DMN connectivity with TMS (8). To identify an individualized DMN map for TMS targeting, a standard DMN template (9) was warped into native space and applied to the participant’s baseline or pre-TMS scan. The participant scan and DMN mask were then warped back into standard MNI space. In each participant, the resultant connectivity maps yielded a correlation cluster in the left posterior inferior parietal lobule. A target was then placed in the averaged center of the left posterior inferior parietal lobule correlation cluster (formed from the overlay of the left posterior inferior parietal lobule clusters derived from the connectivity maps) on the cortical surface using Brainsight neuronavigation software (Rogue Research, Inc.) An individualized TMS target was selected in the left parietal region of the DMN and used as the TMS target for all TMS sessions. Created with BioRender.com.

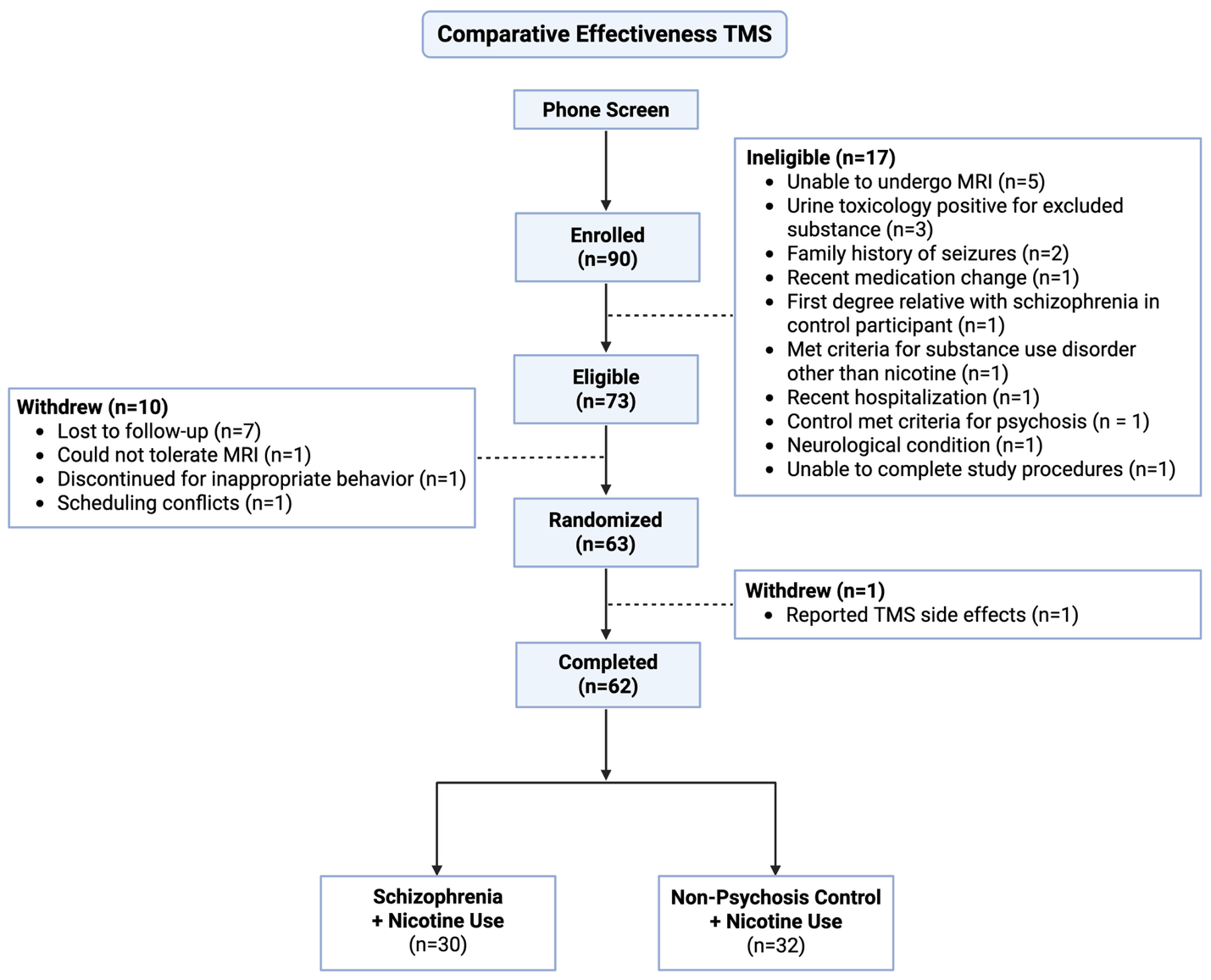
**Supplemental Figure 3. Comparative Effectiveness TMS Consort Diagram.** Ninety participants using nicotine were enrolled. Seventy-three passed screening procedures, and 63 were randomized in our crossover study of individualized, left parietal DMN-targeted TMS compared to L DLPFC-targeted TMS. Sixty-two participants (30 schizophrenia, 32 non-psychosis control) completed the study and provided data for analysis (Figure 1B). Created with BioRender.com.

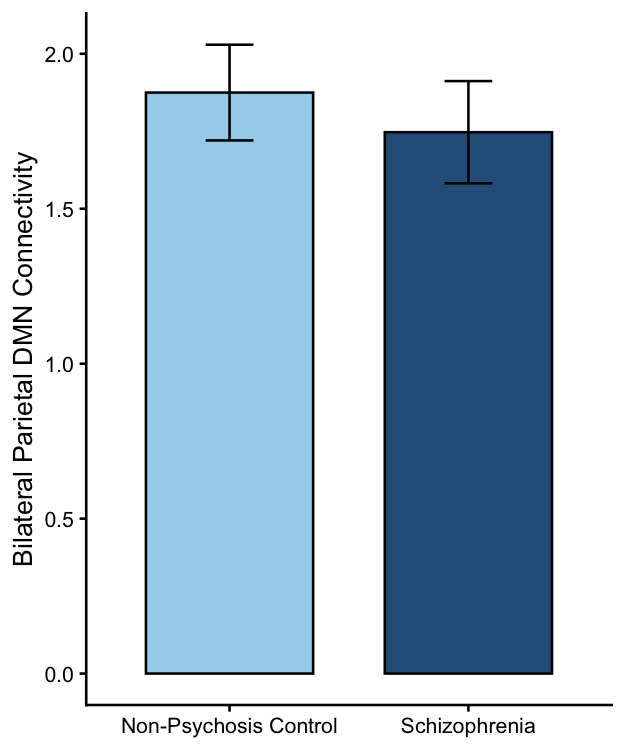

**Supplemental Figure 4. Pre-TMS Bilateral Parietal DMN Connectivity Does Not Differ by Group.** In a linear mixed effects model predicting pre-TMS bilateral parietal DMN connectivity neither age, group, nor their interaction were significant predictors (p>.05).

**Supplemental Table 1. Protocol Optimization TMS Results of Voxel-wise Analysis of Left Lateral Parietal DMN Connectivity and Craving Change**

| **Cluster Index** | **Voxels** | **MAX** | **MAX X (vox)** | **MAX Y (vox)** | **MAX Z (vox)** | **COG X (vox)** | **COG Y (vox)** | **COG Z (vox)** |
| --- | --- | --- | --- | --- | --- | --- | --- | --- |
| 39 | 226 | 1.93 | 24 | 21 | 52 | 25.9 | 28.4 | 57 |
| 38 | 209 | 1.93 | 62 | 79 | 41 | 65.3 | 79.8 | 44.3 |
| 37 | 104 | 1.93 | 68 | 70 | 47 | 67.1 | 66.5 | 50.7 |
| 36 | 74 | 1.93 | 38 | 50 | 18 | 40.9 | 53 | 22 |
| 35 | 51 | 1.93 | 10 | 54 | 22 | 12.9 | 49 | 25.6 |
| 34 | 35 | 1.93 | 14 | 42 | 57 | 14 | 42.2 | 58.6 |
| 33 | 30 | 1.93 | 25 | 82 | 36 | 26.3 | 82.4 | 38.2 |
| 32 | 28 | 1.93 | 61 | 59 | 33 | 60.1 | 61 | 39.2 |
| 31 | 21 | 1.93 | 51 | 47 | 29 | 52.2 | 48.8 | 29.7 |
| 30 | 20 | 1.93 | 58 | 42 | 55 | 60 | 41.5 | 56.3 |
| 29 | 20 | 1.93 | 71 | 38 | 29 | 71.8 | 37.6 | 30.7 |
| 28 | 20 | 1.93 | 31 | 58 | 15 | 29.6 | 57 | 18.2 |
| 27 | 17 | 1.93 | 64 | 84 | 52 | 63.2 | 83.1 | 54.7 |
| 26 | 14 | 1.93 | 49 | 45 | 51 | 47.9 | 44.9 | 52.5 |
| 25 | 11 | 1.93 | 71 | 41 | 59 | 72.8 | 42.7 | 60.9 |
| 24 | 11 | 1.93 | 43 | 57 | 33 | 43.4 | 57 | 34.2 |
| 23 | 11 | 1.93 | 54 | 29 | 17 | 52 | 29 | 19 |
| 22 | 9 | 1.93 | 42 | 46 | 53 | 42.4 | 46.6 | 53.3 |
| 21 | 9 | 1.93 | 26 | 64 | 49 | 25.9 | 64.7 | 49.9 |
| 20 | 8 | 1.93 | 22 | 71 | 41 | 21.5 | 71.9 | 42.2 |
| 19 | 7 | 1.93 | 43 | 32 | 24 | 43.1 | 31.9 | 25.3 |
| 18 | 6 | 1.93 | 22 | 67 | 52 | 20.5 | 68.3 | 53.8 |
| 17 | 6 | 1.93 | 65 | 52 | 17 | 65.3 | 52.2 | 18.2 |
| 16 | 4 | 1.93 | 39 | 55 | 39 | 39.5 | 55 | 39.5 |
| 15 | 3 | 1.93 | 34 | 49 | 38 | 34.7 | 50 | 39 |
| 14 | 3 | 1.93 | 22 | 88 | 36 | 22.3 | 88 | 36.7 |
| 13 | 3 | 1.93 | 39 | 77 | 56 | 39.3 | 77.7 | 56.3 |
| 12 | 3 | 1.93 | 31 | 61 | 32 | 30.7 | 61 | 32.7 |
| 11 | 2 | 1.93 | 36 | 73 | 56 | 36 | 72.5 | 56.5 |
| 10 | 2 | 1.93 | 50 | 30 | 22 | 49.5 | 29.5 | 22.5 |
| 9 | 2 | 1.93 | 42 | 43 | 37 | 42 | 43 | 37.5 |
| 8 | 2 | 1.93 | 60 | 63 | 31 | 60 | 63.5 | 31 |
| 7 | 2 | 1.93 | 73 | 46 | 25 | 73.5 | 46.5 | 25 |
| 6 | 1 | 1.93 | 64 | 63 | 48 | 64 | 63 | 48 |
| 5 | 1 | 1.93 | 66 | 25 | 46 | 66 | 25 | 46 |
| 4 | 1 | 1.93 | 42 | 31 | 22 | 42 | 31 | 22 |
| 3 | 1 | 1.93 | 72 | 60 | 54 | 72 | 60 | 54 |
| 2 | 1 | 1.93 | 23 | 47 | 40 | 23 | 47 | 40 |
| 1 | 1 | 1.93 | 51 | 71 | 24 | 51 | 71 | 24 |

**Supplemental Table 2. Protocol Optimization TMS Mixed Effects Model of Craving Change by Bilateral Parietal DMN Connectivity Change*Age Interaction**

| Predictor | Estimate | SE | Statistic | df | p |
| --- | --- | --- | --- | --- | --- |
| Intercept | 5.74 | 2.00 | 2.87 | 6.57 | **0.026** |
| Bilateral Parietal DMN Connectivity Change | 2.67 | 1.16 | 2.30 | 10.52 | **0.043** |
| Age | -0.11 | 0.05 | -1.97 | 8.14 | 0.084 |
| Bilateral Parietal DMN Connectivity*Age | -0.04 | 0.03 | -1.56 | 14.57 | 0.14 |

**Supplemental Table 3. Comparative Effectiveness TMS Mixed Effects Model of Craving Change by TMS Targets**

| Predictor | Estimate | SE | Statistic | df | p |
| --- | --- | --- | --- | --- | --- |
| Intercept | 0.96 | 0.35 | 2.74 | 109.77 | **0.007** |
| Pre-TMS Craving | -0.47 | 0.07 | -6.86 | 102.20 | **<0.001** |
| TMS Target [DMN] | 0.11 | 0.32 | 0.34 | 60.38 | 0.733 |

**Supplemental Table 4. Comparative Effectiveness TMS Craving Change by TMS Target**

| TMS Target | Mean change | SE | CI | t | df | p |
| --- | --- | --- | --- | --- | --- | --- |
| DLPFC | -0.81 | 0.25 | -1.30, -0.32 | -3.26 | 117.61 | **0.0015** |
| DMN | -0.70 | 0.25 | -1.19, -0.21 | -2.83 | 117.38 | **0.0054** |

**Supplemental Table 5. Comparative Effectiveness TMS Mixed Effects Model of Craving Change by TMS Target*Group Interaction**

| Predictor | Estimate | SE | Statistic | df | p |
| --- | --- | --- | --- | --- | --- |
| Intercept | 0.92 | 0.40 | 2.30 | 112.76 | **0.023** |
| Pre-TMS Craving | -0.47 | 0.07 | -6.74 | 101.59 | **<0.001** |
| TMS Target [DMN] | 0.05 | 0.45 | 0.11 | 59.30 | 0.92 |
| Group [Schizophrenia] | 0.13 | 0.51 | 0.25 | 114.84 | 0.80 |
| TMS Target [DMN] x Group [Schizophrenia] | 0.13 | 0.65 | 0.20 | 59.61 | 0.84 |

**Supplemental Table 6. Comparative Effectiveness TMS Mixed Effects Model of Bilateral Parietal DMN Connectivity Change by TMS Target**

| Predictor | Estimate | SE | Statistic | df | p |
| --- | --- | --- | --- | --- | --- |
| Intercept | 0.90 | 0.34 | 2.64 | 60.00 | **0.011** |
| Pre-TMS Bilateral Parietal DMN Connectivity | -0.63 | 0.09 | -7.27 | 77.51 | **<0.001** |
| TMS Target [DMN] | 0.00 | 0.20 | 0.00 | 50.11 | 1.0 |
| Age | -0.00 | 0.01 | -0.27 | 46.91 | 0.79 |

**Supplemental Table 7. Comparative Effectiveness TMS Bilateral Parietal DMN Connectivity Change by TMS Target**

| TMS Target | Mean change | SE | CI | t | df | p |
| --- | --- | --- | --- | --- | --- | --- |
| DLPFC | -0.33 | 0.15 | -0.62, -0.044 | -2.29 | 95.98 | **0.024** |
| DMN | -0.33 | 0.14 | -0.61, -0.050 | -2.34 | 95.98 | **0.022** |

**Supplemental Table 8. Comparative Effectiveness TMS Mixed Effects Model of Bilateral Parietal DMN Connectivity Change by TMS Target*Group Interaction**

| Predictor | Estimate | SE | Statistic | df | p |
| --- | --- | --- | --- | --- | --- |
| Intercept | 0.60 | 0.39 | 1.55 | 60.99 | 0.127 |
| Pre-TMS Bilateral Parietal DMN Connectivity | -0.60 | 0.08 | -7.11 | 74.50 | **<0.001** |
| TMS Target [DMN] | 0.44 | 0.27 | 1.65 | 47.89 | 0.11 |
| Group [Schizophrenia] | 0.59 | 0.29 | 2.01 | 93.68 | **0.048** |
| Age | -0.00 | 0.01 | -0.24 | 45.98 | 0.81 |
| TMS Target [DMN] x Group [Schizophrenia] | -1.00 | 0.40 | -2.49 | 50.38 | **0.016** |

**Supplemental Table 9. Comparative Effectiveness TMS Bilateral Parietal DMN Connectivity Change by TMS Target*Group**

| TMS Target | Group | Mean change | SE | CI | t | df | p |
| --- | --- | --- | --- | --- | --- | --- | --- |
| DLPFC | Control | -0.58 | 0.19 | -0.96, -0.21 | -3.08 | 93.90 | **0.0027** |
| DLPFC | Schizophrenia | 0.0047 | 0.22 | -0.43, 0.44 | 0.022 | 93.83 | 0.98 |
| DMN | Control | -0.15 | 0.19 | -0.52, 0.23 | -0.77 | 93.90 | 0.44 |
| DMN | Schizophrenia | -0.56 | 0.21 | -0.98, -0.14 | -2.67 | 93.86 | **0.0089** |

**Supplemental Table 10. Comparative Effectiveness TMS Mixed Effects Model of L DLPFC-Right Parietal DMN Connectivity Change by TMS Target**

| Predictor | Estimate | SE | Statistic | df | p |
| --- | --- | --- | --- | --- | --- |
| Intercept | 0.38 | 0.39 | 0.98 | 55.56 | 0.33 |
| Pre-TMS L DLPFC-Right Parietal DMN Connectivity | -0.56 | 0.09 | -6.27 | 84.06 | **<0.001** |
| TMS Target [DMN] | 0.25 | 0.19 | 1.32 | 49.14 | 0.19 |
| Age | -0.00 | 0.01 | -0.28 | 47.61 | 0.78 |

**Supplemental Table 11. Comparative Effectiveness TMS Mixed Effects Model of L DLPFC-Right Parietal DMN Connectivity Change by TMS Target*Group Interaction**

| Predictor | Estimate | SE | Statistic | df | p |
| --- | --- | --- | --- | --- | --- |
| Intercept | 0.34 | 0.45 | 0.75 | 55.39 | 0.46 |
| Pre-TMS L DLPFC-Right Parietal DMN Connectivity | -0.54 | 0.09 | -5.89 | 80.19 | **<0.001** |
| TMS Target [DMN] | 0.43 | 0.26 | 1.67 | 48.54 | 0.10 |
| Group [Schizophrenia] | 0.14 | 0.33 | 0.44 | 87.55 | 0.66 |
| Age | -0.00 | 0.01 | -0.36 | 46.90 | 0.72 |
| TMS Target [DMN] x Group [Schizophrenia] | -0.40 | 0.39 | -1.03 | 50.54 | 0.31 |

**Supplemental Table 12. Comparative Effectiveness TMS Mixed Effects Model of Craving Change by Bilateral Parietal Connectivity Change *Group*Age Interaction**

| Predictor | Estimate | SE | Statistic | df | p |
| --- | --- | --- | --- | --- | --- |
| Intercept | 3.08 | 1.02 | 3.01 | 58.20 | **0.004** |
| Pre-TMS Craving | -0.56 | 0.08 | -7.05 | 84.35 | **<0.001** |
| Pre-TMS Bilateral Parietal DMN Connectivity | -0.06 | 0.21 | -0.28 | 72.31 | 0.78 |
| Bilateral Parietal DMN Connectivity Change | -0.08 | 0.62 | -0.13 | 65.71 | 0.90 |
| Age | -0.05 | 0.02 | -2.15 | 50.88 | **0.036** |
| Group [Schizophrenia] | -3.18 | 1.42 | -2.24 | 48.17 | **0.030** |
| Bilateral Parietal DMN Connectivity Change x Age | 0.00 | 0.02 | 0.04 | 62.02 | 0.97 |
| Bilateral Parietal DMN Connectivity Change x Group [Schizophrenia] | -3.37 | 1.10 | -3.06 | 70.05 | **0.003** |
| Age x Group [Schizophrenia] | 0.11 | 0.04 | 2.54 | 51.09 | **0.014** |
| Bilateral Parietal DMN Connectivity Change x Group [Schizophrenia] x Age | 0.12 | 0.04 | 3.33 | 74.94 | **0.001** |

**Supplemental Table 13. Comparative Effectiveness TMS Craving Change by Bilateral Parietal Connectivity Change*Group*Age**

| Group | Age | Bilateral Parietal Connectivity Change Trend | SE | CI | t | df | p |
| --- | --- | --- | --- | --- | --- | --- | --- |
| Control | 24 | -0.065 | 0.29 | -0.64, 0.51 | -0.22 | 72.5 | 0.82 |
| Schizophrenia | 24 | -0.44 | 0.25 | -0.94, 0.063 | -1.74 | 79.5 | 0.086 |
| Control | 46 | -0.050 | 0.31 | -0.66, 0.56 | -0.16 | 66.1 | 0.87 |
| Schizophrenia | 46 | 2.32 | 0.69 | 0.95, 3.70 | 3.37 | 84.6 | **0.0011** |

**Supplemental Table 14. Comparative Effectiveness TMS Mixed Effects Model of Craving Change by L DLPFC-Right Parietal Connectivity Change *Group*Age Interaction**

| Predictor | Estimate | SE | Statistic | df | p |
| --- | --- | --- | --- | --- | --- |
| Intercept | 3.83 | 1.06 | 3.62 | 55.62 | **0.001** |
| Pre-TMS Craving | -0.68 | 0.08 | -8.06 | 87.79 | **<0.001** |
| Pre-TMS L DLPFC-Right Parietal DMN Connectivity | -0.23 | 0.20 | -1.16 | 71.62 | 0.25 |
| L DLPFC-Right Parietal DMN Connectivity Change | 0.07 | 0.56 | 0.13 | 83.27 | 0.90 |
| Age | -0.06 | 0.03 | -2.24 | 48.87 | **0.030** |
| Group [Schizophrenia] | -2.30 | 1.52 | -1.51 | 45.62 | 0.14 |
| L DLPFC-Right Parietal DMN Connectivity Change x Age | -0.00 | 0.01 | -0.25 | 73.76 | 0.81 |
| L DLPFC-Right Parietal DMN Connectivity Change x Group [Schizophrenia] | 0.37 | 0.95 | 0.39 | 80.74 | 0.70 |
| Age x Group [Schizophrenia] | 0.07 | 0.05 | 1.52 | 47.03 | 0.14 |
| L DLPFC-Right Parietal DMN Connectivity Change x Group [Schizophrenia] x Age | -0.04 | 0.03 | -1.20 | 79.30 | 0.23 |

**Supplemental Table 15. Comparative Effectiveness TMS Pre-TMS Bilateral Parietal Connectivity Association with Group*Age**

| Predictor | Estimate | SE | Statistic | df | p |
| --- | --- | --- | --- | --- | --- |
| Intercept | 1.75 | 0.56 | 3.11 | 52.80 | **0.003** |
| Age | 0.00 | 0.01 | 0.27 | 52.90 | 0.79 |
| Group [Schizophrenia] | -0.33 | 0.86 | -0.38 | 55.14 | 0.71 |
| Age x Group [Schizophrenia] | 0.01 | 0.02 | 0.28 | 56.98 | 0.78 |
